# Efficacy and Safety of TurmXTRA^®^ 60N in Delayed-onset Muscle Soreness in Healthy, Recreationally Active Subjects: a Randomized, Double-blind, Placebo-controlled Trial

**DOI:** 10.1101/2022.05.10.22274831

**Authors:** Shefali Thanawala, Rajat Shah, Vasu Karlapudi, Prabakaran Desomayanandam, Arun Bhuvanendran

## Abstract

**Background:** Delayed onset of muscle soreness (DOMS) and its physiological consequences has an important influence on the individual’s adherence to the exercise routine.

**Objective:** This study aimed to evaluate the efficacy, safety, and tolerability of TurmXTRA^®^ 60N (WDTE60N) on DOMS in comparison with placebo in recreationally active healthy subjects.

**Methods:** The present study was a randomized, double-blind, placebo-controlled parallel group study. Thirty healthy and recreationally active subjects (average age: 28.23±4.20 years) were randomized to receive WDTE60N (WDTE60N group; n=15) or placebo (placebo group; n=15). Study treatments were initiated 29 days prior to the eccentric exercise and continued for 4 days after the exercise. Primary endpoint was the change in pain intensity measured by the visual analog scale (VAS) at the end of study treatment (at 96 hours after eccentric exercise) from baseline (measured immediately after exercise).

**Results:** The VAS score indicated that subjects from the WDTE60N group reported significantly less pain after eccentric exercise compared to placebo group (AUC_0-96h_: 286.8±46.7 vs. 460±40.5, respectively; p<0.0001). Wellbeing status, assessed using the adapted version of Hooper & MacKinnon questionnaire, calculated as individual and cumulative scores of the domains - fatigue, mood, general muscle soreness, sleep quality and stress demonstrated significant improvement in all domains as well as in overall wellbeing in WDTE60N group as compared to placebo group (p<0.0001). Serum lactate dehydrogenase (LDH) was significantly lower in the WDTE60N group compared to placebo group (AUC_0-96h_: 23623.7±2532.0 vs. 26138.6±3669.5, respectively; p=0.0446).

**Conclusion:** The WDTE60N intake before and after eccentric exercise significantly reduced subjective perception of muscle soreness and serum LDH activity, and increased psychological wellbeing after eccentric exercise in recreationally active subjects.

## INTRODUCTION

Delayed onset muscle soreness (DOMS) is a type of ultrastructural muscle damage caused by high-intensity eccentric muscle contractions or unaccustomed forms of exercise.(1) The symptoms include impaired muscular force capacities, muscle stiffness, aching pain, and/or muscular tenderness with some altered biomechanics to the adjacent joints.(2-4) It usually begins 6-12 hours after the completion of exercise, peaks at 48-72 hours, and slowly resolves in 5-7 days.(5) However, it is one of the responsible elements for compromised sportive performances. Thus, recovery is of utmost importance as it allows for a quicker return to the next bout of the exercise and perhaps leads to improved adaptations and performance in subsequent exercise bouts. Several dietary supplements are found valuable in enhancing muscular adaptations to exercise, improving brain performance, decreasing DOMS or pain, reducing injury severity and enhancing recovery from injury. Notably, these dietary supplements do not produce ergogenic effects but assist the athletes to train and/or compete more effectively without performance impairments.(6).

Curcumin is a natural polyphenolic substance extracted from rhizomes of the turmeric plant. It is the major constituent (∼77% of total curcuminoids) present in the turmeric rhizome. It gives a distinctive yellow color to the spice turmeric. Owing to its numerous physiological effects, it has been used since the ancient times in traditional Indian and Chinese Medicine. Evidence demonstrates its beneficial effects in treating cancer, cardiovascular health, arthritis and Alzheimer’s dementia that is indicative of its anticarcinogenic, anti-inflammatory, antioxidant and neuroprotective properties.(7-10) Curcumin produces analgesic effects on acute and chronic pain by de-sensitizing the transient receptor potential vanilloid 1, an ion channel responsible for pain sensation, thereby reducing pain sensitivity.(11) It also decreases post-exercise inflammation by regulating Nuclear factor kappa B (NF-□B) and nuclear factor erythroid 2–related factor 2 (Nrf2) pathways that ultimately reduces pain sensitivity and DOMS.(12) It is hypothesized that curcumin, being a free radical scavenger, reduces secondary muscle damage after exercise. Taking into considerations the proven benefits of curcumin, the International Olympic Committee consensus have classified it as a nutritional supplement that may improve training capacity, recovery, muscle soreness, and injury management.(13)

One of the factors that limits the widespread use of curcumin is its poor oral bioavailability, however, this can be overcome by administering very large doses of approximately 6–8 g turmeric powder or 1500–2000 mg turmeric extract standardized to 95% for gaining optimal effects. However, long-term administration of such high doses of curcumin is discouraged in terms of gastrointestinal side effects (intestinal disturbances and urticaria) and poor compliance.(14) TurmXTRA^®^ 60N (WDTE60N; of Nutriventia – Inventia Healthcare Ltd.), a novel formulation of natural, water-dispersible turmeric extract containing 60% natural curcuminoids, is developed using a patented technology to overcome the aforementioned limitations of curcumin. Pre-clinical pharmacokinetic study using Sprague Dawley rats demonstrated ten-fold higher bioavailability with WDTE60N compared to standard turmeric extract 95%.(15) These results have been reciprocated in a comparative pharmacokinetic study that evaluated pharmacokinetics of WDTE60N compared to standard turmeric extract 95% in healthy subjects.(16) Findings of this study indicate that water dispersibility of WDTE60N aids in absorption of active moieties resulting in higher levels of plasma-free curcumin, total curcumin and total curcuminoids compared to standard turmeric extract 95% when subjects were administered a 10-fold lower quantity of curcuminoids in WDTE60N. Recently, we assessed efficacy of WDTE60N in alleviating symptoms of chronic knee pain.(17) Results of the randomized study confirmed significantly greater pain reduction among those receiving WDTE60N as compared to those receiving placebo as evident by significant reduction in VAS score (−1.5 ± 0.7 vs -0.6 ± 0.8, p < 0.0001). Further, the results for time taken for 80-m fast-paced walk test and 9-step stair-climb test with WDTE60N intake were also satisfactory.

Accordingly, this randomized controlled trial was designed to assess efficacy, safety and tolerability of TurmXTRA^®^ 60N (WDTE60N) compared to placebo on DOMS in recreationally active subjects. We hypothesized that consistent use of single daily dose of 250 mg TurmXTRA^®^ 60N during pre- and post-period of eccentric exercise reduces DOMS and muscle damage.

## MATERIALS AND METHODS

### Study design

The present randomized, double-blind, placebo-controlled parallel group study was conducted between 29 Dec 2021 and 07 Feb 2022 after receiving approval from an institutional ethics committee of the study center (ACE Independent Ethics Committee, registration detail: ECR/141/Indt/KA/2013/RR-19). The study was conducted in accordance with the pertinent requirements of the Declaration of Helsinki (Brazil, October 2013), Good Clinical Practices for Clinical Research in India 2005, New Drugs and Clinical Trials Rules 2019, ICH E6 (R2), Guidance on Good Clinical Practice, and with ICMR’s National Ethical Guidelines for Biomedical and Health Research Involving Human Participants-2017. All subjects were thoroughly informed about the study prior their enrollment and before obtaining their signed informed consent forms. This study was registered with the Clinical Trial Registry of India (registration no. CTRI/2021/12/038892).

### Study population

The present study was conducted in 30 healthy and recreationally active individuals. The subjects were included if they met following inclusion criteria: 1) Male and female subjects with ages 18 to 35 years having body mass index of 18 to 30 kg/m^2^; 2) Healthy, moderately active [regular aerobic exercise for at least 150 min per week for past 3 months], non-smoker subjects without any known musculoskeletal pathology; 3) Subjects vaccinated (two doses) for Coronavirus disease 2019 (COVID-19); 4) Female subjects of childbearing potential were included only if they agreed to use medically acceptable form of birth control; 5) Subjects willing to sign the informed consent and comply with the study procedure; 6) Subjects willing to abstain from using analgesics/non-steroidal anti-inflammatory drugs during the treatment period; and 7) Subjects whose screening test results were within the normal range or were not considered clinically significant by the Principal Investigator. However, the subjects were excluded from the study if: 1) Female subjects were pregnant, breastfeeding or planning to get pregnant; 2) Subjects had COVID-19 symptoms; 3) Subject had known allergy to any compound present in the study product; 4) Subject was regularly involved in strength training; 5) Subjects had a history or frequently uses over-the-counter non-steroidal anti-inflammatory drugs and/or analgesic agents; 6) Subjects with high alcohol intake (>2 standard drinks per day) or use recreational drugs (cocaine, methamphetamine, marijuana, etc.) or have nicotine/caffeine dependence; 7) Subjects with a history/presence of chronic illness including but not limited to psychiatric (psychosis, major depressive disorder or other clinically significant psychiatric disorder), cardiovascular, pulmonary, neurological, gastrointestinal, hepatic, renal, metabolic, and ophthalmologic disorders or with human immunodeficiency virus infection and acquired immune deficiency syndrome or those taking regular prescription pharmacological agents; 8) Subjects who participated in any other trials involving investigational or marketed products within 30 days prior to the screening visit; 9) Subjects who underwent surgery or trauma affecting the knee in the past; 10) Subjects with a history or presence of any neurological or muscular disorders that may affect muscle strength; 11) Subjects regularly taking multivitamins/herbal and/or turmeric supplements/ any other wellness product.

### Study procedure

The study was conducted at Arogyavardhini Ayurvedic Centre, Bangalore, Karnataka, India. All subjects underwent a screening evaluation 7 days prior administering study treatments. Screening evaluation included anthropometry measurements, vital sign assessment, physical examination, medical and medication history (including history of substance abuse and or addiction to drugs, cigarette smoking and alcohol intake, any concomitant medication usage), urine pregnancy test for female subjects with child-bearing potential, electrocardiogram, laboratory investigations for liver function, renal function, hematology, fasting blood glucose and human immunodeficiency virus infection. All subjects were given a COVID-19 screening questionnaire and those who fulfilled all the eligibility criteria were randomly allocated to two treatment groups using blocked randomization method on Day 1, WDTE60N group (n=15) or placebo group (n=15). The randomization schedule was generated using SAS version 9.4 (SAS Institute Inc., USA) by an independent statistician and it was made available to the study personnel responsible for packaging, labeling, and blinding of the investigational product. The subjects were enrolled by the study investigator. Blinding was performed via computer generated process. The study team statistician was responsible for the blinding process. Randomization numbers were pre-printed as stickers and were affixed on each bottle that was the only identity to separate one kit from the other. Investigator assigned treatment kits to the enrolled subjects as per the randomization schedule. Subjects were instructed to administer study treatment, either a capsule containing 250 mg water dispersible turmeric extract (WDTE60N) or placebo, once daily, orally with adequate water after breakfast. The subjects were required to administer the study products for 33 days (pre-exercise period of 29 days and post-exercise period of 4 days). They were also required to maintain a subject diary that would include details regarding their daily intake of study products and concomitant medication (if taken). Dosing compliance was assessed by reviewing subject diary at each visit. Subjects performed eccentric exercise (squatting) on Day 29. Squatting involves quadriceps muscles undergoing eccentric contractions in the downward phase of the movement. According to eccentric exercise protocol, each subject was required to perform one set of squats (measured as 15 squat cycles) in one minute and continue to perform the same for 15 minutes to complete 225 squat cycles. However, subjects were allowed to stop the exercise if they were unable to continue when the neuromuscular system could no longer produce adequate force to proceed or when the investigator felt that the exercise was sufficient to induce DOMS.

### Study endpoints

Primary efficacy endpoint was improvement in pain intensity from baseline (immediately after eccentric exercise) to end of the treatment (at 96 hours after eccentric exercise).

Pain was assessed by asking the question, ‘What is the intensity of your current pain?’ and the response was recorded on a visual analogue scale (VAS) that had a 10 cm line with “no pain” on one end and “worst imaginable pain” on the other end. The VAS score was measured immediately before, immediately after (baseline), and at 12, 24, 48, 72 and 96 hours after eccentric exercise.

Secondary efficacy endpoints of the study were as follows:

1 Improvement in the domains of fatigue, sleep quality, general muscle soreness, stress level, mood and overall wellbeing from baseline (immediately after exercise) to the end of treatment (at 96 hours after exercise) as evaluated by adapted version of Hooper & MacKinnon questionnaire. The adapted version was used for ease of administration in the study.

Self-reported ratings of well-being may provide an efficient means of monitoring both overtraining and recovery. Hooper & MacKinnon questionnaire quantified and qualified the well-being relative to fatigue, sleep quality, general muscle soreness, stress and mood.(18) This self-reported well-being questionnaire is often used as a repeated measure to investigate changes over time in individuals, or as a general indicator of well-being across the group to understand general patterns.(19) Further, it is found sensitive to variations within and between weeks.(20) The Hooper & MacKinnon questionnaire was later updated by MacLean in 2014.(21) The adapted version of the Hooper & MacKinnon questionnaire provides a subjective assessment of the impact of DOMS in the domains of fatigue, sleep quality, general muscle soreness, stress level and mood on a scale of 1-5, with higher scores denoting the better results (Table 2).

In the present study, the well-being of the subjects was assessed using the adapted version of Hooper & MacKinnon questionnaire immediately before, immediately after (baseline), and at 12, 24, 48, 72 and 96 hours after eccentric exercise.

2. Change in serum creatine kinase (CK) and lactate dehydrogenase (LDH) from baseline (immediately before eccentric exercise) to end of the treatment (at 96 hours after eccentric exercise)

The muscle damage markers (serum CK and serum LDH activity) were measured immediately before (baseline), and at 24, 48, 72 and 96 hours after eccentric exercise.

3. Subject’s and physician’s global assessment of therapy at the end of treatment (at 96 hours after eccentric exercise) (Figure 1 A).

The safety assessment of the study products was based on the adverse events reported, changes in vital parameters and laboratory investigations (complete blood count, liver function test, renal function test) as well as general wellbeing.

**Figure 1:**
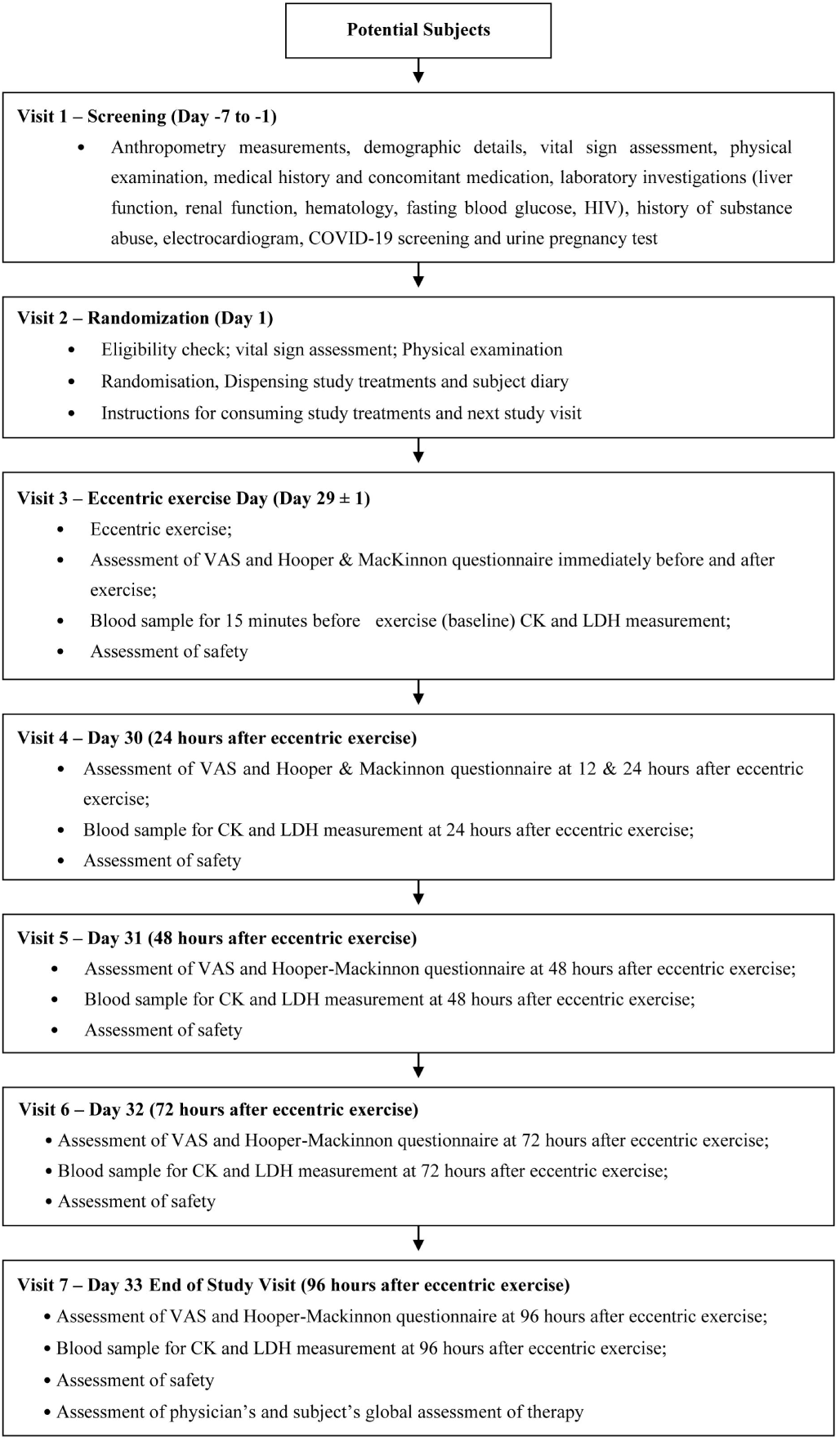
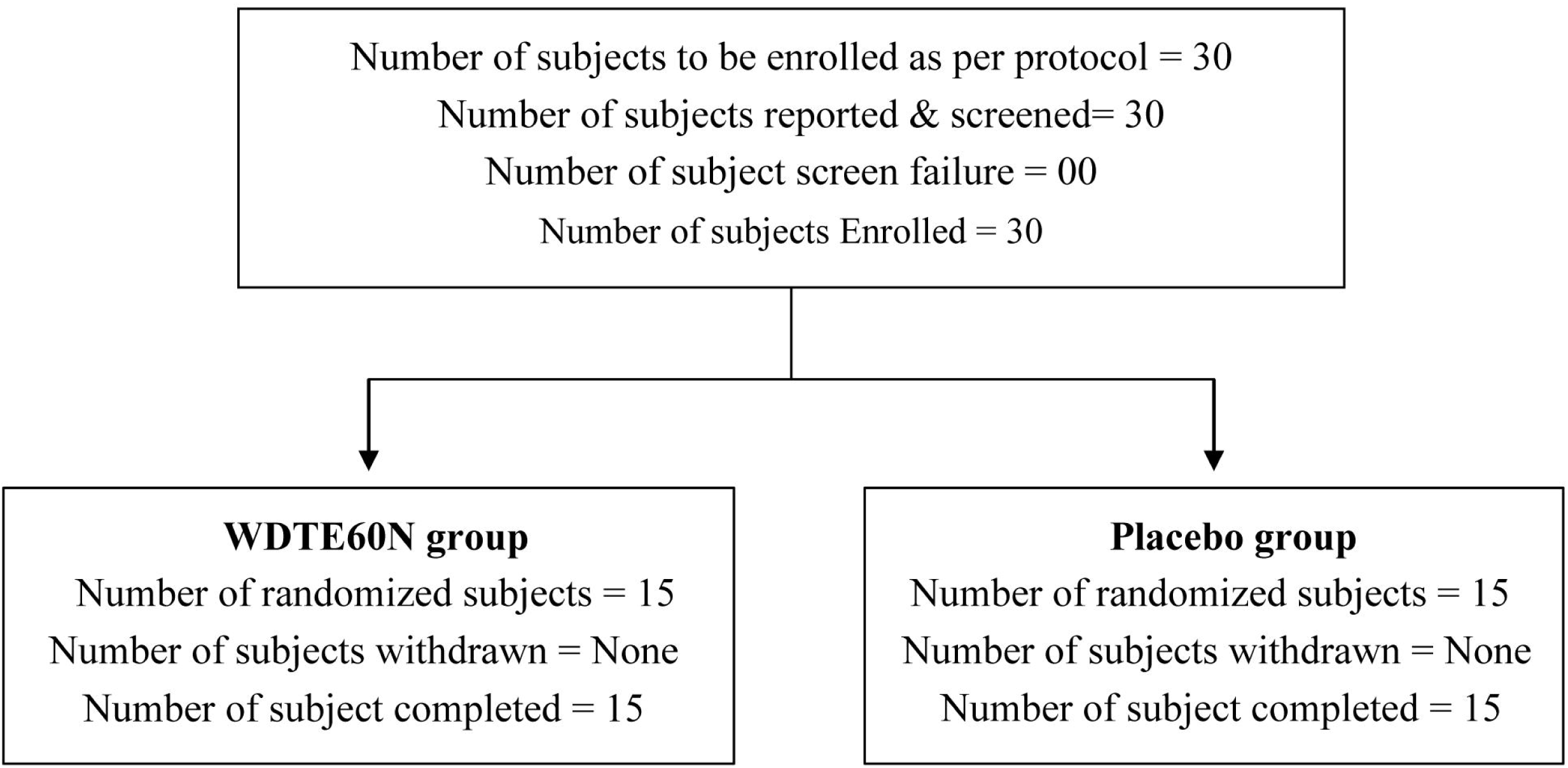
Subject disposition and flow of the study Figure 1: (A) Flow of the study Figure 1 (B): Subject disposition

### Statistical analysis

Present study was an exploratory study and the sample size was calculated based on the published literature. Considering the power of 80%, alpha of 0.05, anticipated mean VAS score for WDTE60N group and placebo group of 2.88 and 3.36, respectively, sample size of 28 subjects was required and after including 10% of dropout, the total sample size was estimated to be 30 subjects.

Subjects were included in the Per Protocol population if they completed the study treatment without any major protocol deviations with treatment compliance of at least ≥ 80% were considered. The results were presented as mean ± standard deviation for continuous variables and count and percentages for categorical variables. Kolmogorov– Smirnov test was performed to assess normality distribution of the data. The unpaired t-test was used for intra treatment group comparison depending upon normality distribution of continuous. The study involves evaluation of efficacy of test product in DOMS- a condition in which the pathogenesis is known to start - attain peak -decline over a period of time after performing unaccustomed eccentric muscular activity, hence assessment of efficacy of the product cannot be significantly established at any single time point after the eccentric exercise. Therefore, area under curve (AUC) was utilized to describe the span of the efficacy parameters over time after the DOMS induction. A p-value<0.05 was considered statistically significant. Statistical analysis was performed using SAS version 9.4 (SAS Institute Inc., USA).

## RESULTS

From 29 Dec 2021 to 06 Jan 2022, 30 recreationally active subjects (mean age: 28.23 ± 4.20 years) were screened and enrolled in the study. Of them, 21subjects were males, 09 females. The enrolled subjects were randomly allocated to the WDTE60N group (n=15) and placebo group (n=15). All subjects completed the study (**Figure 1 B**). Demographic and baseline clinical characteristics of the study population are shown in **Table 1**.

**Table 1:** Demographics and baseline clinical characteristics of the study population (n=30 subjects)

| **Characteristics** | **Overall**<br>**(N=30)** | **WDTE60N group**<br>**(n=15)** | **Placebo group**<br>**(n=15)** |
| --- | --- | --- | --- |
| Age (years), Mean±SD | 28.23±4.20 | 29.7±3.6 | 26.8±4.3 |
| Number of male subjects | 21 | 11 | 10 |
| Number of female subjects | 09 | 04 | 05 |
| Weight (kg), Mean ± SD | 59.98±3.21 | 60.27±3.58 | 59.69±2.89 |
| Height (cm), Mean ± SD | 170.84±6.37 | 170.73±6.37 | 170.95±6.58 |
| Body mass index (kg/m<sup>2</sup>),<br>Mean ± SD | 20.59±1.16 | 20.72±1.28 | 20.45±1.04 |

**Table 2:** Hooper & MacKinnon questionnaire (adapted version) used to assess improvement in the domains of fatigue, sleep quality, general muscle soreness, stress level, mood and overall wellbeing

| **Parameters** | **Score** |  |  |  |  |
| --- | --- | --- | --- | --- | --- |
|  | **5** | **4** | **3** | **2** | **1** |
| **Fatigue** | Very Fresh | Fresh | Normal | More tired than normal | Always tired |
| **Sleep quality** | Very Restful | Good | Difficulty falling asleep | Restless sleep | Insomnia |
| **General Muscle Soreness** | Feeling great | Feeling good | Normal | Increasing in soreness / tightness | Very sore |
| **Stress Levels** | Very relaxed | Relaxed | Normal | Feeling stressed | Highly stressed |
| **Mood** | Very positive mood | A generally good mood | Less interested in other and/or activities than usual | Less interested in other and/or activities than usual | Highly annoyed/ irritable/ down |

Both the groups were found statistically insignificant and comparable for efficacy parameters (VAS score, all domains of well-being status, serum CK and LDH concentration) when evaluated immediately before exercise

### Visual analogue scale

Both treatment groups had statistically insignificant and comparable VAS score immediately after eccentric exercise (baseline). The VAS score significantly lowered in the WDTE60N group compared to the placebo group at 12 hours (4.0±0.7 vs. 4.5±0.6; p=0.0320), 24 hours (5.0±0.8 vs. 5.9±0.5; p=0.0010), 48 hours (3.3±0.7 vs. 5.5±0.6; p<0.0001), 72 hours (1.9±0.7 vs. 4.5±0.6; p<0.0001) and 96 hours (0.7±0.5 vs. 3.3±0.5, p<0.0001) after exercise (**Figure 2 A**). Mean AUC from immediately after exercise to the end of the study (AUC_0-96h_) was significantly lower in the WDTE60N group compared to placebo group (286.8±46.7 vs. 460±40.5; p<0.0001) (**Supplementary Figure 1**).

**Figure 2.**
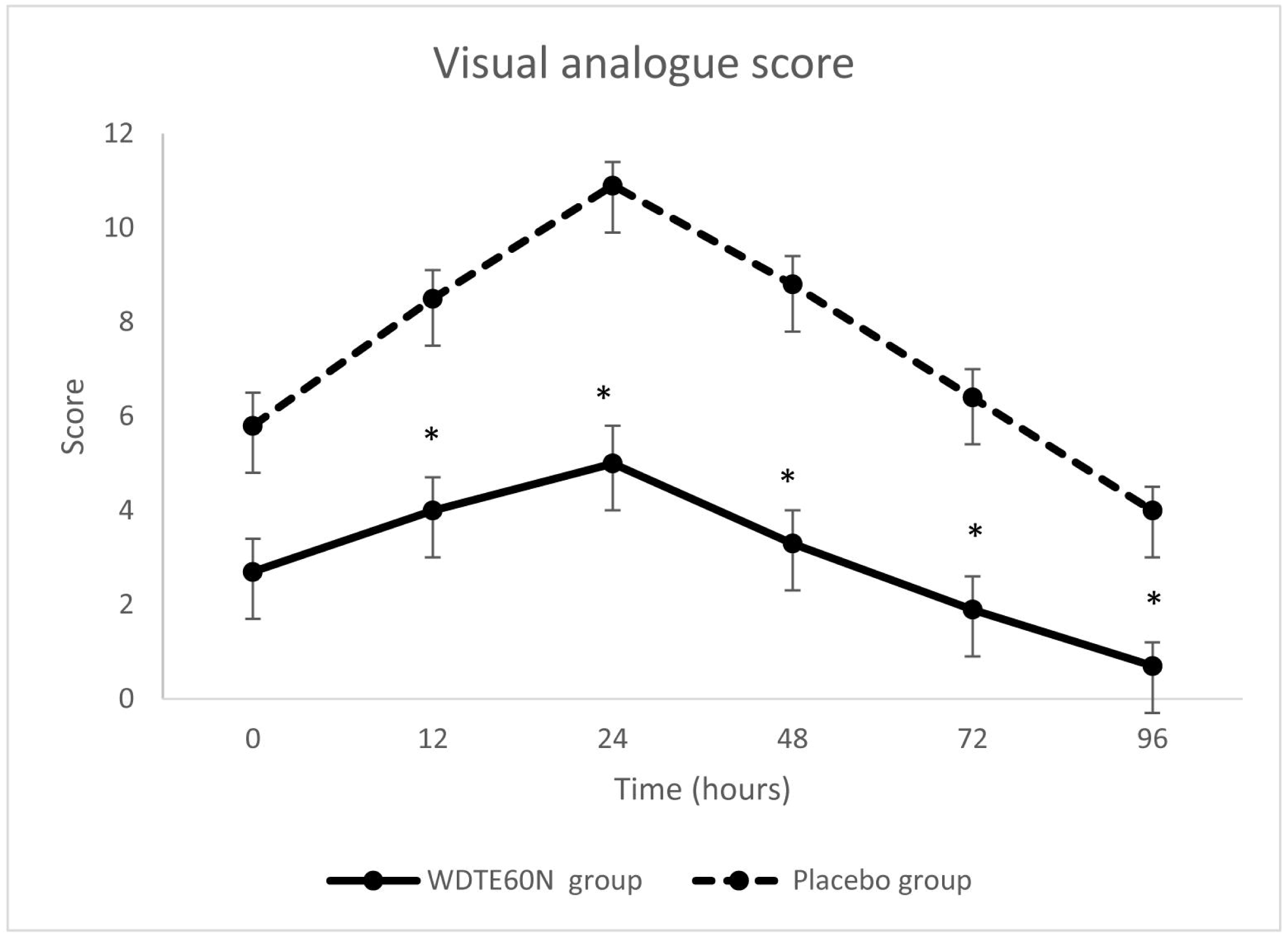
Changes in VAS score immediately after (0 h) and 12-96 hours (12, 24, 48, 72 and 96h) after eccentric exercise in the subjects receiving WDTE60N and placebo; Values are presented as mean±SD (n=15 for each group); *p<0.05

### Assessment of wellbeing status using adapted version of Hooper & MacKinnon questionnaire

Both treatment groups were comparable at immediately before and after eccentric exercise as evident by no statistically significant difference for all five domains and overall well-being at baseline (p>0.05) (**Figure 3**). However, there was significant improvement in all five domains at different time points as follows:

**Figure 3:**
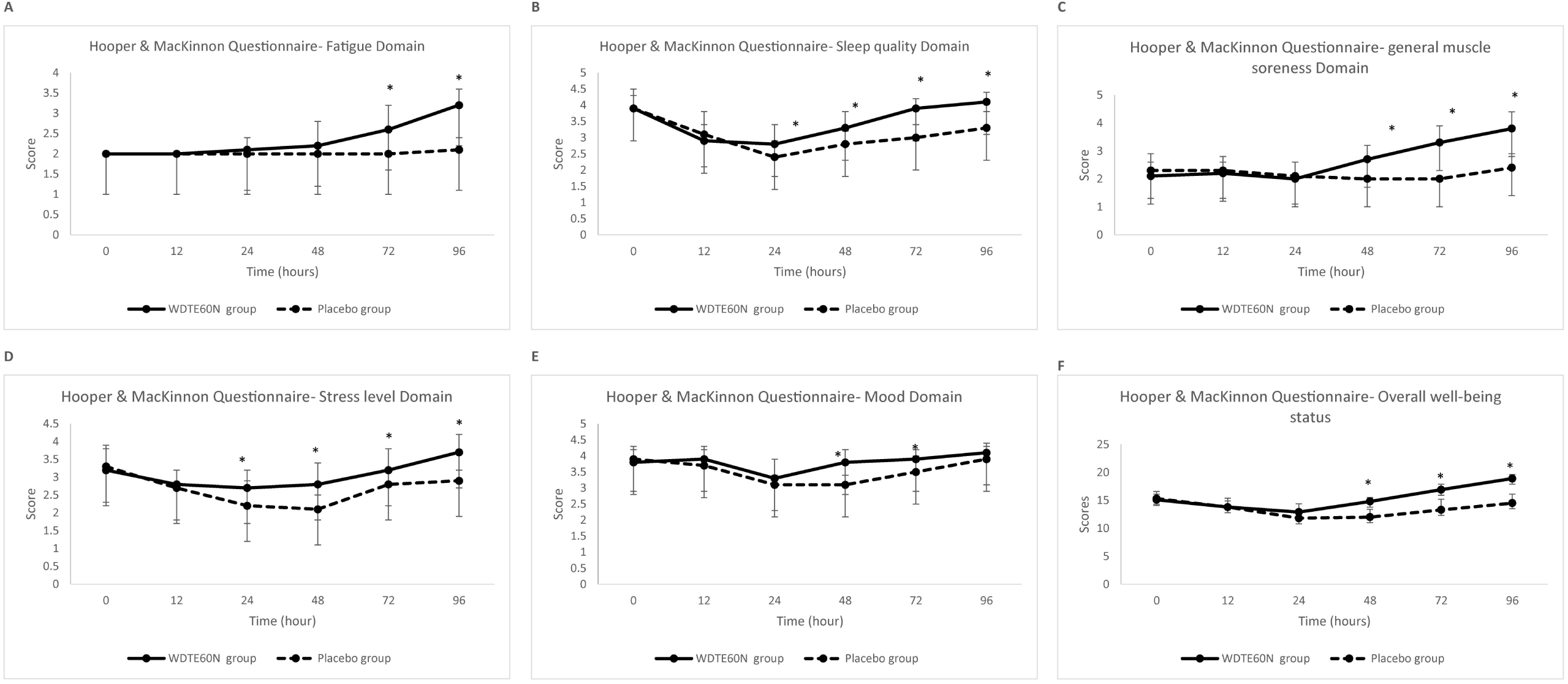
Changes in wellbeing status assessed using adapted version of Hooper & MacKinnon questionaire immediately after (0 h) and 12-–96 hours (12, 24, 48, 72 and 96h) after eccentric exercise in the subjects receiving WDTE60N and placebo; Values are presented as mean±SD (n=15 for each group); *p<0.05 Figure 3 (A): Changes in fatigue domain assessed using adapted version of Hooper & MacKinnon questionnaire immediately after and 12-–96 hours (12, 24, 48, 72 and 96h) after eccentric exercise in the subjects receiving WDTE60N and placebo; Values are presented as mean±SD (n=15 for each group); *p<0.05 Figure 3 (B): Changes in sleep quality domain assessed using adapted version of Hooper & MacKinnon questionnaire immediately after (0 h) and 12-–96 hours (12, 24, 48, 72 and 96h) after eccentric exercise in the subjects receiving WDTE60N and placebo; Values are presented as mean±SD (n=15 for each group); *p<0.05 Figure 3 (C): Changes in general muscle soreness domain assessed using adapted version of Hooper & MacKinnon questionnaire immediately after (0 h) and 12-–96 hours (12, 24, 48, 72 and 96h) after eccentric exercise in the subjects receving WDTE60N and placebo; Values are presented as mean±SD (n=15 for each group); *p<0.05 Figure 3 (D): Changes in stress level domain assessed using adapted version of Hooper & MacKinnon questionnaire immediately after (0 h) and 12-–96 hours (12, 24, 48, 72 and 96h) after eccentric exercise in the subjects receiving WDTE60N and placebo; Values are presented as mean±SD (n=15 for each group); *p<0.05 Figure 3 (E): Changes in mood domain assessed using adapted version of Hooper & MacKinnon questionnaire immediately before, immediately after (0 h) and 12-–96 hours (12, 24, 48, 72 and 96h) after eccentric exercise in the subjects receiving WDTE60N and placebo; Values are presented as mean±SD (n=15 for each group); *p<0.05 Figure 3 (F): Changes in overall wellbeing status assessed using adapted version of Hooper & MacKinnon questionnaire immediately after (0 h) and 12-–96 hours (12, 24, 48, 72 and 96h) after eccentric exercise in the subjects receiving WDTE60N and placebo; Values are presented as mean±SD (n=15 for each group); *p<0.05

### Fatigue

The fatigue score was significantly higher at 72 hours (2.6±0.6 vs. 2.0±0; p=0.0025) and 96 hours (3.2±0.4 vs. 2.1±0.3; p<0.0001) after the exercise in WDTE60N group as compared to placebo group.

### Sleep Quality

The WDTE60N group reported significantly higher score at 24 hours (2.8±0.6 vs. 2.4±0.5; p=0.0499), 48 hours (3.3±0.5 vs. 2.8±0.4; p=0.0032), 72 hours (3.9±0.3 vs. 3.0±0.4; p<0.0001) and 96 hours (± vs. ±; p<0.0001) after exercise.

### General muscle soreness

There was significantly higher general muscle soreness score at 48 hours (2.7±0.5 vs. 2.0±0.0; p=0.0001), 72 hours (3.3±0.6 vs. 2.0±0.0; p<0.0001) and 96 hours (3.8±0.6 vs. 2.4±0.5; p<0.0001) after exercise in WDTE60N group as compared to placebo group.

### Stress Level

The scores for stress levels were significantly higher in WDTE60N group as compared to placebo group at 24 hours (2.7±0.5 vs. 2.2±0.7; p=0.0388), 48 hours (2.8±0.6 vs. 2.1±0.4; p=0.0005), 72 hours (3.2±0.6 vs. 2.8±0.4; p=0.0345) and 96 hours (3.7±0.5 vs. 2.9±0.3; p<0.0001) after exercise.

### Mood Level

In comparison with placebo group, the WDTE60N group showed significantly higher mood scores at 48 hours (3.8±0.4 vs. 3.1±0.3; p<0.0001) and 72 hours (3.9±0.3 vs. 3.5±0.5; p=0.0051) after exercise.

### Overall well-being

The overall well-being scores were significantly higher at 48 hours (14.8±1.4 vs. 12.0±0.7; p<0.0001) and 72 hours (16.9±1.9 vs. 13.3±1.0; p<0.0001) and 96 hours (18.9±1.6 vs. 14.5±0.7; p<0.0001) after exercise.

Corresponding mean AUC for the aforementioned domains was also significantly higher in WDTE60N group compared to the placebo group (**Supplementary Figure 2**).

### Serum creatine kinase concentration

There was no statistically significant difference in the pre-exercise serum CK concentration between the WDTE60N group and placebo group (115.3±26.5 IU/L and 170.0±136.6 IU/L; p=0.1483). Serum CK concentration peaked at 48 hours (152.9±52.4 IU/L) but decreased at 72 hours (125.9±41.2 IU/L) and 96 hours (138±53.6 IU/L) after eccentric exercise in the WDTE60N group. Contrastingly, in the placebo group, there was a decrease in serum CK concentration initially at 24 hours and 48 hours followed by an increase at 72 hours and 96 hours after exercise. Decrease in the mean AUC_0-96h_ serum CK was observed in the WDTE60N group compared to placebo group. However, the difference between both treatment groups at different time points and in AUCs was statistically insignificant.

### Serum LDH concentration

Pre-exercise serum LDH concentration was 262.3±151.3 U/L and 253.8±72.0 U/L for the WDTE60N and placebo group, respectively, and the difference was not statistically significant. Serum LDH concentration peaked at 48 hours, which decreased gradually at 72 hours and 96 hours after exercise in both groups (**Figure 4)**. It was significantly lowered in subjects in the WDTE60N group at 72 hours (223.2±38.5 vs. 298.5±95.0; p=0.0106) and 96 hours (206.6±25.4 U/L vs. 277.8±98.6 U/L; p=0.0156) compared to those in placebo group. Statistically significant decrease in the mean AUC_0-96h_ serum LDH concentration was observed in the WDTE60N group compared to placebo group (23623.7±2532.0 vs. 26138.6±3669.5; p=0.0446) **(Supplementary Figure 3)**.

**Figure 4:**
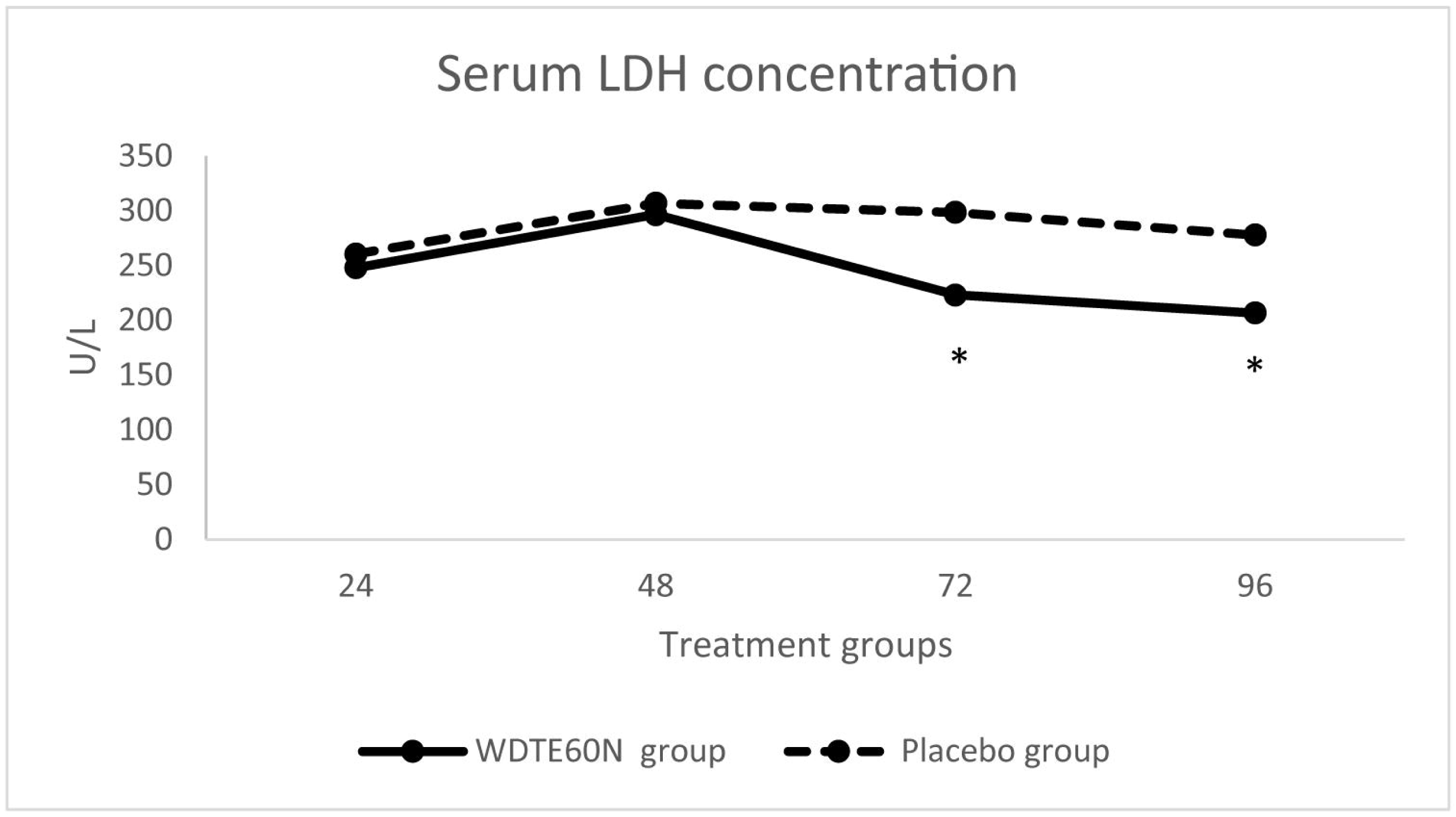
Changes in serum LDH activity after 24-96 hours (24, 48, 72 and 96 hours) of eccentric exercise in the subjects receiving WDTE60N and placebo; Values are presented as mean ± SD (n=15 subjects in WDTE60N-group; n=14 subjects in placebo-group); *p<0.05

### Determination of the subject’s and physician’s global assessment of therapy

The subject’s global assessment of therapy in both WDTE60N and placebo groups were 4.00±0.0 and 2.00±0.0, respectively (p<0.05). Similarly, the physician’s global assessment of therapy showed significantly higher scores for WDTE60N group compared to placebo group (4.07±0.26 vs. 2.40±0.51; p<0.0001).

### Safety and tolerability of the study treatments

No incidence of serious adverse events was reported during the entire study, and the study treatments were well tolerated by the subjects. There were no clinically significant changes in vital signs, routine examinations of hematology, liver function test and renal function test in the study subjects of both treatment arms.

## DISCUSSION

The study outcomes demonstrated the effects of the WDTE60N on muscle soreness and muscle damage markers after intensive eccentric exercise in recreationally active subjects compared to placebo. Main findings of our study included: (1) the pain intensity after eccentric exercise was significantly lowered among the subjects receiving WDTE60N compared to those receiving placebo at 12, 24, 48 and 96 hours after eccentric exercise; 2) significantly higher improvement in fatigue, mood, sleep quality, generalised muscle soreness, stress and overall wellbeing (assessed by the adapted Hooper & MacKinnon questionnaire) was noted in subjects receiving WDTE60N than those on placebo; 3) Significantly decreased serum LDH concentrations but not serum CK concentration with consistent use of WDTE60N supplement before and after eccentric exercise; and 4) significantly higher ratings for WDTE60N in the subject’s and physician’s global assessment of therapy. Further, the study treatments were well tolerated by all subjects, and no serious adverse events were reported during the study period.

Several studies demonstrated efficacy of curcumin in reducing muscle soreness or pain associated with exercise-induced muscle damage.(22-25) The comparison of acute pain relieving effects of curcumin (400 mg), nimesulide and acetaminophen showed comparable analgesic activity of curcumin with that of acetaminophen. Further, curcumin also possesses better gastric tolerability than nimesulide.(26) In this study, assessment of pain intensity using the VAS score demonstrated significantly lower pain in subjects receiving WDTE60N throughout the post-exercise period compared to those receiving placebo. Notably, this lower pain was evident (p<0.05) within 12 hours of eccentric exercise thus indicating faster recovery. Similarly, assessment of wellbeing status using the adapted version of Hooper & MacKinnon questionnaire confirmed less muscle soreness in subjects receiving WDTE60N (p<0.05). Moreover, overall wellbeing and other aspects such as fatigue, mood, sleep quality and stress also significantly improved after exercise in subjects who received WDTE60N than those administered placebo. The impact of WDTE60N on stress levels was as in accordance with previous study. Sciberras et al. reported higher number of subjects with “better than usual” scores in the subjective assessment of psychological stress during training day when the subjects were receiving curcumin supplementation prior to the training.(27) Further, the subjects who feel the pain and the physicians treating it may have different assessments and perception of the pain, which could affect the evaluation of treatment efficacy. Thus, this needs to be evaluated from both subject’s and physician’s perspectives. In the current study, both, the subject’s rating and the physician’s rating for global assessment of therapy were significantly higher for WDTE60N. This observation confirms that both subjects and physicians perceived WDTE60N as an effective treatment option to counter physiological effects of DOMS. The pain alleviating efficacy of WDTE60N has also been demonstrated for management of chronic knee pain. The study also demonstrated improvements in joint performance and mobility without any significant safety concern following WDTE60N administration once daily for 3 months.(17) Thus, the results of the previous and present studies reflect that WDTE60N could have beneficial effects on the entire musculoskeletal system with a single daily dose of one capsule.

Serum CK concentration is found to be increased after eccentric exercise as it is well distributed in muscle tissue and released into the circulation according to a loss of sarcolemmal integrity as a result of mechanical stress of eccentric exercise or metabolic causes like glycogen depletion. Similarly, increased serum LDH concentrations indicates cell damage as it is present in the cytoplasm and catalyzes the reversible process of pyruvate to lactate under anaerobic conditions. Thus, an increase in serum CK or LDH activities are indirect markers of muscle damage. In this study, serum CK and LDH concentrations increased and attained highest levels at 48 hours following exercise in both treatment groups thus indicating DOMS to be a clinical condition with peak clinical implications within 72 hours of eccentric exercise. Serum LDH concentration significantly lowered in the WDTE60N group compared to the placebo-group at 72 and 96 hours after exercise. Similarly, after exercise, reduced serum CK activity was found in the subjects receiving WDTE60N as compared to those on placebo (p>0.05). Results from several studies have warranted the need to evaluate the effect of curcumin supplements on muscle damage markers to determine the correlation of frequent or higher dosing with decrease in muscle damage markers. A study that evaluated effects of a curcumin and piperine combination on the recovery kinetics after exercise-induced muscle damage, reported no effect of treatment on serum CK concentration.(28) Similarly, Drobnic et al. determined the effects of curcumin (initiated 48 hours prior exercise and continued for 24 hours after exercise) compared with placebo on exercise-induced DOMS where the authors reported no difference in CK concentration between both groups but noted significantly lowered levels of interleukin-8 in those given curcumin after exercise (29).

While evaluating safety, the present study reveals that WDTE60N intake for 33 days neither caused any adverse effects nor reported any change in vital or laboratory parameters. These observations confirm that WDTE60N is safe and tolerable for human consumption. In contrast, non-steroidal anti-inflammatory drugs, a commonly used treatment strategy to relieve muscle soreness associated with DOMS by athletes, has several side effects.

Briefly, intense muscle soreness decreases strength and muscle power ultimately leading the athletes to perform the next bout of exercise at a higher intensity than to which they are normally habituated.(30) This increased intensity due to damaged and weakened muscle fibers (alterations in muscle sequencing and recruitment patterns) following eccentric exercise will likely increase the risk for further injury if premature return to exercise is attempted.(31) Based on the results of this present study, we believe that WDTE60N intake is advantageous to athletes as it decreases subjective and objective parameters of muscle soreness. Further, it also reduces the level of stress that may improve the performance of athletes.

### Limitations

We acknowledge the limitations of this study. The present study did not determine the effect of therapy in athletes who trained and competed regularly. Further studies can be planned to evaluate efficacy of the WDTE60N in athletes.

## CONCLUSION

This study demonstrated that WDTE60N administration, both prior and after eccentric exercise, facilitates quicker reduction of pain and return to exercise routine and may allow the subjects to undergo higher intensity exercises as it reduces post-exercise pain, improves the wellbeing status including muscle soreness and decreases lactate accumulation as demonstrated by significantly lowered serum LDH activity without any major safety concern in recreationally active subjects.

## Data Availability

All data produced in the present work are contained in the manuscript

## DECLARATIONS

### Data sharing statement

All data generated or analyzed during this study are included in this published article along with its supplementary information file.

### Conflicts of Interest

Rajat Shah and Shefali Thanawala are employees of Nutriventia – Inventia Healthcare Ltd. Prabakaran Desomaya nandam and Arun Bhuvanendran are employees of In Vitro Research Solutions (iVRS) Pvt Ltd. Authors do not have any other conflicts of interest to declare.

## Acknowledgments

The study was sponsored by Nutriventia – Inventia Healthcare Ltd., India. The authors would like to acknowledge CBCC Global Research team for their writing and editorial support in the development of this manuscript.

## Author Contributions

Shefali Thanawala, and Rajat Shah conceived the study. Prabakaran D and Arun B conducted the study, undertook data collection, and performed the data analyses. Shefali Thanawala interpreted the results with assistance from Vasu Karlapudi, and Prabakaran B. Arun B prepared the first draft. All authors have read and agreed to the published version of the manuscript.

## Funding

The study was funded by Nutriventia – Inventia Healthcare Ltd.

## Notes

### Clinical Trial

CTRI/2021/12/038892

### Author Declarations

Institutional ethics committee of the study center (ACE Independent Ethics Committee, registration detail: ECR/141/Indt/KA/2013/RR-19) gave approval for this work.

