## Supplementary figures and images for "Efficacy and Safety of TurmXTRA^®^ 60N in Delayed-onset Muscle Soreness in Healthy, Recreationally Active Subjects: a Randomized, Double-blind, Placebo-controlled Trial"

### Supplementary Figure 1

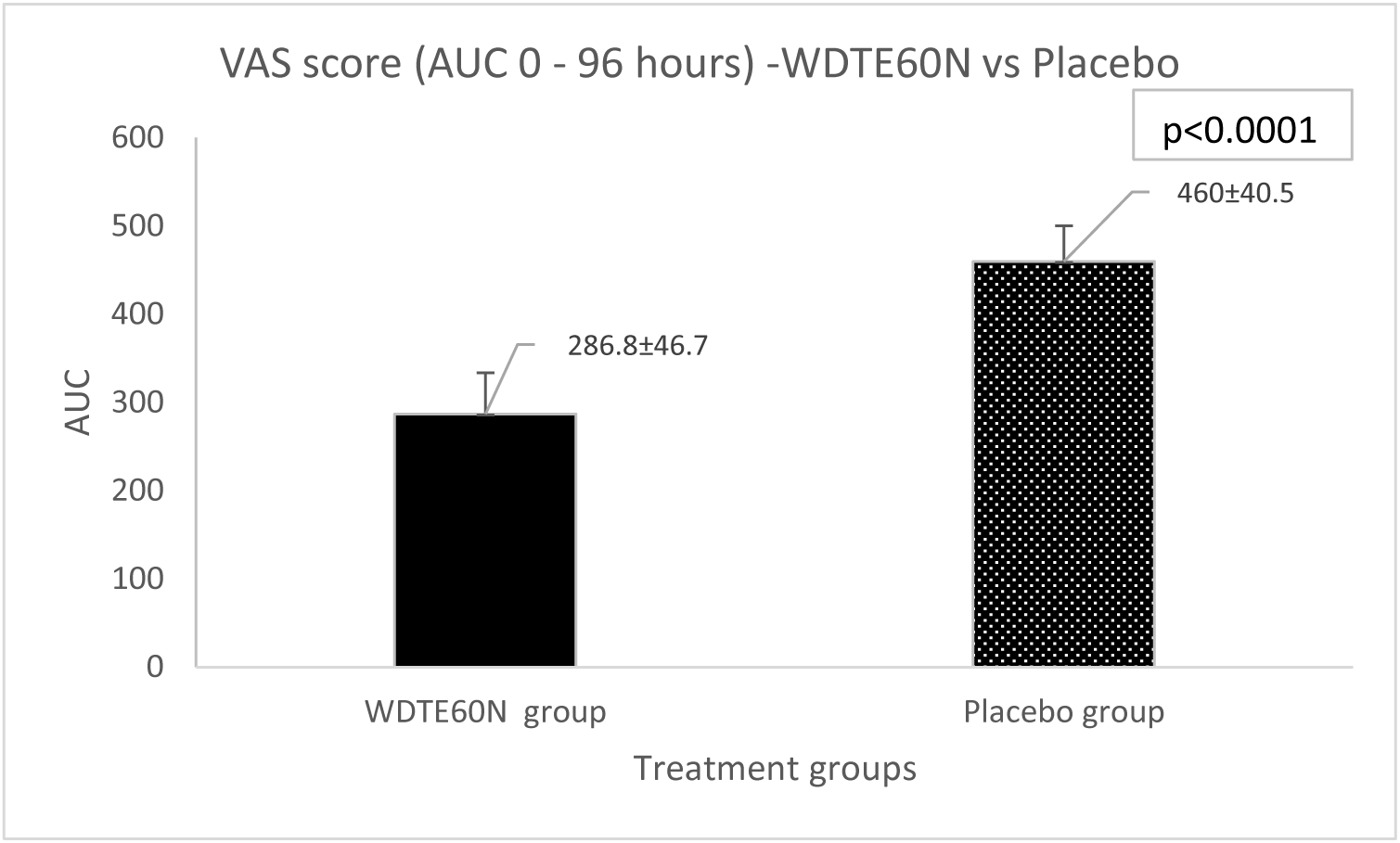

### Supplementary Figure 2

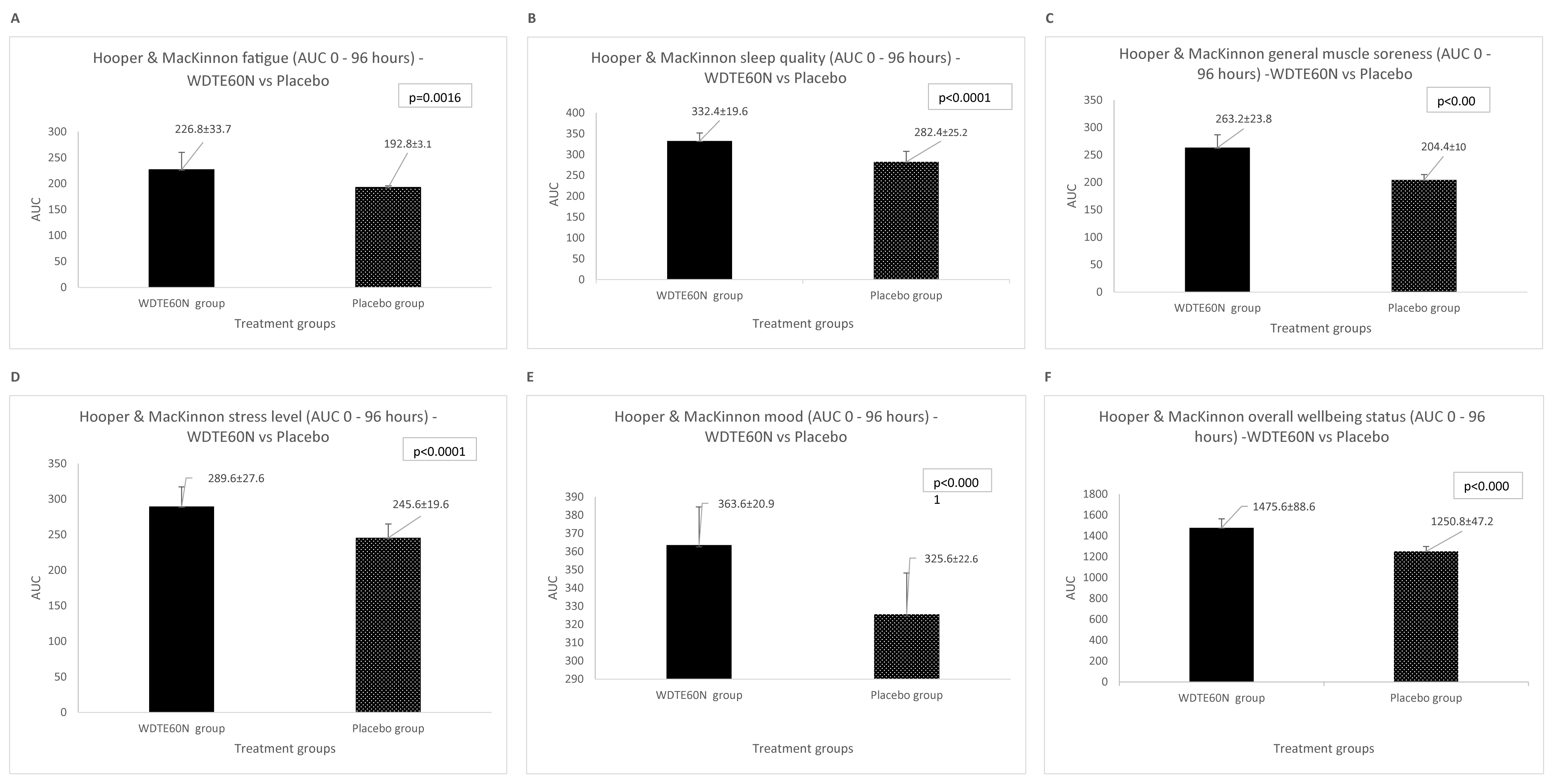

### Supplementary Figure 3

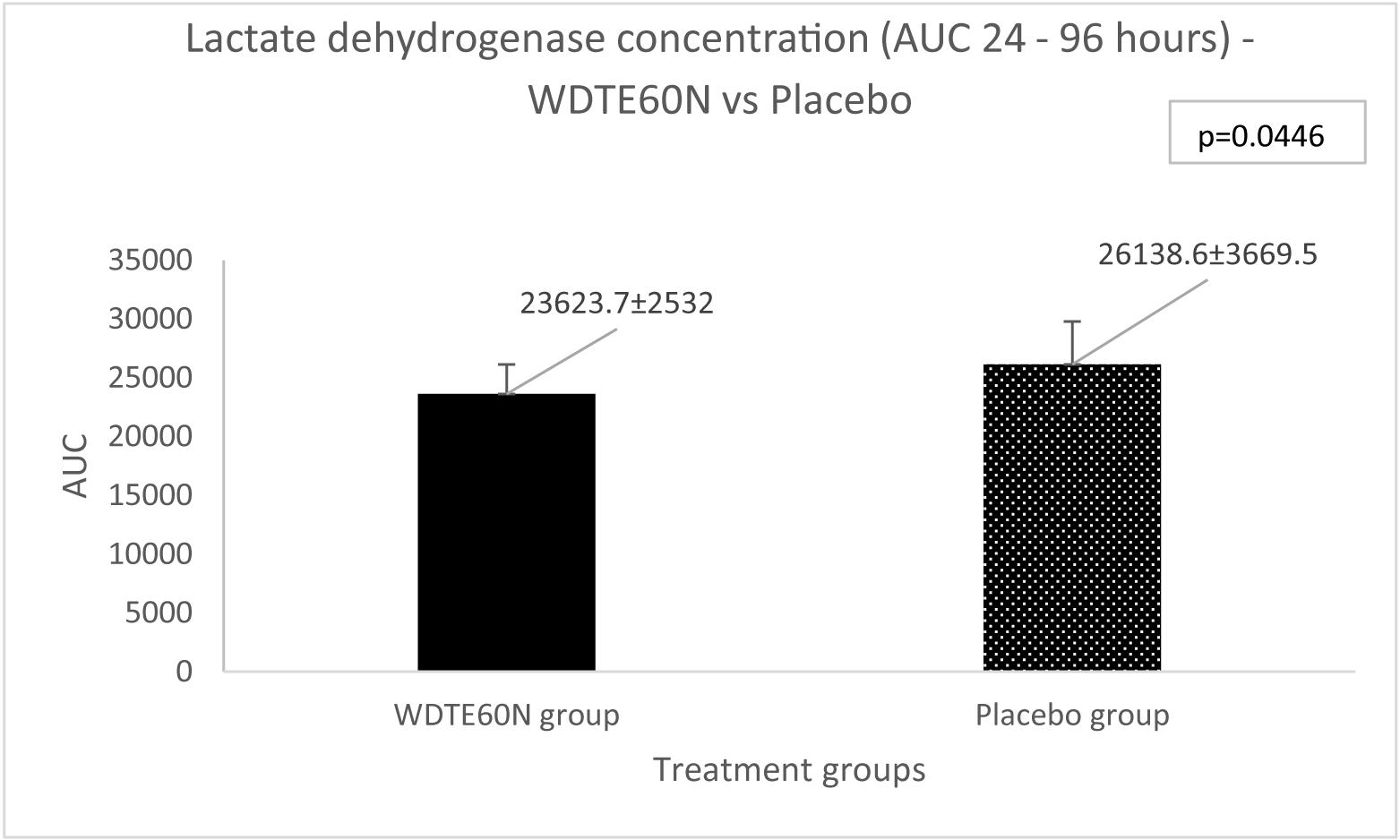
